# Development of a cognitive behavioral therapy intervention in Pujehun District, Sierra Leone among pregnant women

**DOI:** 10.1101/2024.12.21.24319013

**Authors:** Ashleen Lee, Aminata Koroma, Joshua A Duncan, Abdulai Jawo Bah, Eliza Kleban, Kristin Kohlmann, Jumeika Lopez-Arteaga, Kevin Stephenson, Mark J Manary

## Abstract

This report outlines the development of a novel therapeutic intervention to address perinatal depression (PND) in Pujehun District, a rural area in Sierra Leone marked by high illiteracy rates and socio-economic challenges. PND poses significant health risks for both mothers and their children yet remains largely unaddressed due to stigma and limited resources.

In collaboration with the Mental Health Coalition – Sierra Leone, Project Peanut Butter established the first cognitive behavioral therapy (CBT) service in the district for women with PND. This process began with the creation of the first mental health counseling service in the district for women with postnatal depression, along with the selection and training of female counselors. A culturally adapted CBT curriculum was developed, designed to empower participants to reintegrate into their communities. Additionally, a novel screening tool was introduced to effectively diagnose PND in pregnant women in low-literacy settings.

This model serves as a replicable framework for similar initiatives in high illiteracy contexts, with the goal of improving maternal mental health and ultimately infant development.

## Introduction

Depressive disorder affects approximately 322 million individuals globally (WHO, 2017). Depression is particularly pronounced in sub-Saharan Africa (SSA), where neuropsychiatric disorders account for nearly 10% of disease burden (Tomlinson et al., 2009). Depression is 50% more prevalent in women than men, and the perinatal period is an especially vulnerable time to develop mood disorders (WHO, 2023). Perinatal depression (PND) is the occurrence of a depressive episode during pregnancy and/or following childbirth (Dagher et al., 2021). Despite the presence of effective interventions for mental health disorders, over 75% of individuals in low- and middle-income countries do not have access to mental health care (WHO, 2023). Barriers to receiving mental health care include inadequate investment in mental health services, shortage of trained healthcare professionals, and persistent stigma surrounding mental disorders.

PND poses substantial health risks for both the mother and child (Dagher et al., 2021). Antenatal depression is associated with a higher incidence of prematurity, low birth weight, and maternal complications such as pre-eclampsia, placental abnormalities, and spontaneous abortion. Postpartum depression perturbs maternal-infant bonding, adaptation to the mothering/caretaking role, maternal quality of life, and the children’s emotional, cognitive, and behavioral development. Risk factors for PND include a history of depression, history of physical or sexual abuse, unplanned/unwanted pregnancy, stressful life events, lack of social and financial support, intimate partner violence and medical complications of pregnancy. Low socioeconomic status, lack of social support, and bearing children during adolescence have also been shown to increase women’s risk of developing PND after delivery (O’Connor et al., 2019). A high adherence to healthy dietary patterns is inversely associated with depression (Psaltopoulou et al., 2013).

In Sierra Leone, where women are regularly exposed to many of these stressors and lack of resources, PND is common. PND and anxiety affect about 23% of pregnant women in Africa (Endomba et al., 2021). Our provisionary work with a mental health service undertaken in Pujehun District June 2022 – October 2022 estimated that 10% of malnourished women met diagnostic criteria for depression. According to the Sierra Leone Demographic and Health Survey, 61% of married women say they have experienced physical, sexual, or emotional spousal violence. Due to the culture condoning gender-based violence, which includes shame, stigma, and economic concerns regarding the subject, most incidents go unreported (M’Cormack-Hale & Akua Amoah Twum, 2022). Further, Sierra Leone has faced multiple traumatic societal events in recent history including a civil war between the years of 1991 and 2002, the Ebola Virus Disease outbreak in 2014 that disrupted post-war efforts to rebuild infrastructures, and most recently the COVID-19 pandemic, leaving Sierra Leone as one of the poorest countries in the world and ranked 182 out of 189 on the Human Development Index of 2020 (Lane, 2021).

Lower literacy is associated with greater mental health disorders (Hunn et al., 2023). Sierra Leone has one of the highest illiteracy rates in the world (World Population Review, 2024). In this context, literacy is defined as being able to read and write in English, Arabic, Krio, Mende, or Temne, two local languages. Based on previous data attained in 2019 in Pujehun District, only 12.9% of females classified as literate, indicated by completion of primary school or above.

Cognitive behavioral therapy (CBT) is based on the paradigm that thoughts can create feelings and that feelings drive behavior (Figure 1). CBT is a form of psychotherapy in which patients work with a mental health counselor to identify and alter maladaptive thinking, and which improves emotional response and behavior going forward (Mayo Clinic, 2019). The aim of CBT is to help participants respond to challenging situations in more productive manners.

**Figure 1.**
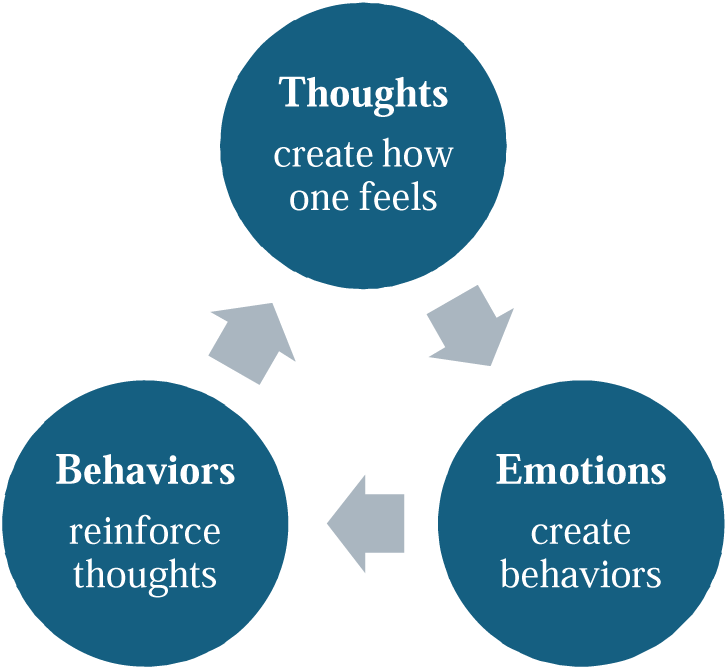
Foundation of CBT.

A meta-analysis of the implementation of CBT found that signs and symptoms of depression resolved more quickly than with no intervention (Hofmann et al., 2012). A culturally relevant form of CBT accounts for common philosophical assumptions, including acculturation, beliefs regarding health and illness, help-seeking behaviors, and cultural orientation towards psychotherapy (Rathod et al., 2019). CBT in its current form cannot be used in communities with low literacy rates, as its implementation requires that participants complete reading and writing exercises.

Despite the numerous risk factors that women in Sierra Leone face, there is a paucity of mental health resources available. The Mental Health Coalition (MHC)– Sierra Leone was established in 2011, at a time when there was only one working psychiatrist in the country and had only recently trained its first psychiatric nurse (*Mental Health Coalition – Sierra Leone | MHIN*, 2014). This lack of resources has left gaps in addressing mental health needs, particularly for perinatal women.

Given these unmet needs, Project Peanut Butter (PPB) created a small-scale perinatal mental health counseling service in Pujehun District. After the establishment of this service, a culturally appropriate perinatal depression screening tool and CBT curriculum were developed. The formation of this novel service is described in more detail.

## Methods

### Setting

This mental health outreach project was established in rural Pujehun District in the Southern Province of Sierra Leone and is deeply rooted in the local context of the district, which reflects both available resources and cultural practices. Meals regularly consist of rice or cassava, served with sauces or soups made from cassava leaf, ground nut, or other available vegetables or fish. Traditional cooking methods such as boiling, steaming, or frying are commonly used. Homes are typically constructed of mud with thatch or metal roofs without indoor plumbing or electricity. Sierra Leonean culture is patriarchal, and it is common for a man to take multiple wives through customary marriage. Family sizes can be large as it is common for every wife to have 3 to 4 children. Due to the rural setting of the district, it is common for individuals to walk several miles on bush paths to access facilities such as health centers in larger villages. The other common mode of transportation is motor bikes. The population is predominantly Muslim and mainly belongs to the Mende ethnic group (ReliefWeb, 2016).

The project development and conduct of the subsequent clinical trail was approved by the Sierra Leone Ethics and Research Committee, Government of Sierra Leone, Freetown, Sierra Leone and the Washington University Human Studies Committee, St. Louis, MO USA.

### Project Peanut Butter (PPB)

PPB is a nonprofit organization locally registered in Sierra Leone that aims to advance the treatment of malnutrition. PPB provides necessary nutritional and medical support to women and children suffering from acute malnutrition in sub-Saharan Africa through the production and distribution of effective Ready to Use Therapeutic Foods (RUTF) and Ready to Use Supplemental Foods. PPB operates out of mobile clinics throughout sub-Saharan Africa to assess women and children for malnutrition and provide treatment. PPB is responsible for provision of products and services for malnutrition treatment and prevention in Pujehun District.

### Mental Health Coalition (MHC) - Sierra Leone

The MHC-SL was established in August 2011 to address ongoing mental health challenges in Sierra Leone. Its mission is to empower stakeholders to advocate for their needs, thereby elevating the profile of mental health in the country. As the primary reference point for mental health in Sierra Leone, the MHC-SL has facilitated significant initiatives such as the launch of the National Mental Health Policy and Strategic Plan, integration of mental health into the national Poverty Reduction Strategy Paper, and establishment of the national Mental Health Steering Committee at the Ministry of Health and Sanitation (MoHS). The MHC-SL has positioned itself as a vital resource during crises, such as the Ebola outbreak, becoming a key partner of the MoHS and the Ministry of Social Welfare, Gender & Children’s Affairs (Mental Health Innovation Network, 2014).

The MHC-SL was consulted by PPB to create the first counseling service and mental health intervention to mothers suffering from postpartum depression in Pujehun District. This initiative involved the development of training materials, identification and training of counselors, and providing ongoing support to ensure the effectiveness and quality of the service. The MHC-SL also provided pre-existing tools to aid in the screening process of the novel mental health counseling service.

### Selection, training and activities of Counseling Staff

Twenty candidates for the counseling staff were selected from local female residents in Pujehun District that self-identified as having an interest in initiating local women’s groups. Candidates were chosen for their ability to communicate and show empathy toward local participants, rather than for meeting any formal education requirements. Candidates completed a series of training sessions and evaluations on the identification and management of postpartum depression facilitated by members of the MHC-SL. This curriculum was based on a manual developed by the MHC-SL that was the result of consultation with international and local partners including clinicians, social workers, advocates, and tutors. The manual was pre-tested and validated for use. Five counsellors were subsequently chosen by PPB based on the following criteria:

1. Gender: female
2. Location: within Pujehun proximity
3. Experience: had encountered or seen others who may have been affected by PND
4. Knowledge: excelled in training based on feedback; strong understanding of local dialect
5. Strong social quotient: ability to start, grow, and maintain conversations at all levels regardless of social and/or economic status

The group of counselors chosen for the team consisted of three social workers who had graduated from university, one nurse and one advocate who had not graduated.

MHC-SL staff oversee supervision of the counseling staff in Pujehun District to build skills and problem solve with counselors. Two MHC-SL experts visit Pujehun monthly and accompany counselors during clinic sessions throughout the week. During these visits, MHC-SL staff can assess counselor performance and speak with participants privately to evaluate the quality of counseling services. Monthly visits also allow MHC-SL staff to enhance counselors’ training in the field as needed.

#### Initial Activities of Counseling Staff

This mental health intervention began at five prenatal clinics within Pujehun District in 2022 and consisted of three key phases: screening, counselling, and integration.

Screening was conducted at clinic sites, where counselors were able to effectively use three pre-existing tools developed by the MHC-SL to screen for PND, measure function, and measure stress levels in pregnant women. The screening phase revealed the following common conditions among those with postpartum depression:

- Early motherhood (teenage years)
- Abandonment by partner due to pregnancy
- Withdrawal from education while peers remained enrolled
- Physical abuse by partner during pregnancy
- “Psychological torture” such as partner infidelity, neglect, or lack of financial support
- Abandonment by parents due to shame

The counselling phase aimed to help participants identify sources of distress by introducing the concept of Specific, Measurable, Attainable, Realistic, Timely (SMART) goals. This approach was designed to allow participants to address their own concerns and alleviate depressive symptoms. Counselling consisted of seven 45-minute sessions, with the fifth session dedicated to group therapy. The following is an example of one’s participant’s progression through counselling:

The participant, a young mother, expressed stress and fear regarding financial stability after her spouse left her while pregnant. The counselor encouraged her to consider potential solutions, leading the participant to realize she needed to find a new source of income. With this goal in mind, the counselor introduced the concept of SMART goals. Recognizing her own skillset in baking, the participant was able to construct a plan to profit from this talent. Her sessions progressed as follows:

1. Identify the cause of distress; consider baking as a potential solution
2. Discuss logistics of where and when to gather ingredients, items to bake, where to sell goods, etc. through the lens of SMART goals
3. “Assigned” to prepare goods to be sold next session for counselors
4. Home visit: counselors checked on the progress participant made at home
5. Group session
6. Progress update

a. Participant decided to sell goods at clinic site
7. Update and check in regarding participant’s mental and emotional state

Group therapy sessions provided a safe and empowering space for participants to share their stories with one another, fostering mutual support and connection. Counselors were also able to build confidence in their skills through the structure of group therapy, as they were each assigned a role during the sessions. This consisted of leading the group prayer, breathing exercises, ice breakers, discussion, and singing.

Once a participant completed their seven counselling sessions, they were re-assessed with a pictorial screening tool developed by PPB. This tool was designed to determine whether the participant was eligible to graduate from the program. If the assessment indicated that the participant was not ready to graduate, three additional sessions were added to their program. Counseling continued until subsequent assessment confirmed readiness to graduate. The numbers served during the first 6 months of the screening and counselling phases are shown in Table 1.

**Table 1.** Numbers Served during first 6 months of counselling service.

| **Clinic** | **Number Screened** | **Number Counsellled** |
| --- | --- | --- |
| Bandajuma | 125 | 82 |
| Potoru | 106 | 77 |
| Pujehun | 146 | 36 |
| Sahn Malen | 243 | 98 |
| Zimmi | 120 | 46 |
| **Total** | **740** | **339** |

The following is an excerpt from a PPB staff member during the implementation of this mental health service:

“In the initial weeks of launching the Mental Health Program (MHP), the team visited five clinics, building rapport with the chiefs by explaining the program’s purpose, discussing suitable spaces for counseling sessions, and gathering essential clinic information, such as the number of recent pregnant women, check-up schedules, and optimal days for counseling.
Each counselor carried general consent forms, screening tools, and pens. As women waited for their check-ups, counselors approached them to explain the program and gauge their interest in joining. After obtaining consent, counselors used the screening tool to assess depression. If a woman screened positive, she was assigned an identification number for attendance tracking and provided her with an appointment card for counseling sessions. The challenges faced, included heavy rain, scheduling conflicts with cultural activities, low attendance, and difficulty contacting participants due to unreliable addresses and phone numbers.
Once counseling sessions began, each counselor allocated 45 minutes for one-on-one interactions. Counselors sought outdoor areas to ensure confidentiality. When confronted with lack of participant motivation, SMART goals were employed to enable participants to set achievable targets. We also discussed using a ruler question to gauge participants’ willingness to change behaviors. As participants accomplished their weekly goals, their confidence and mood improved. For example, one of the participants at Zimmi Clinic’s goal was to become financially independent and support her child. Their counselor helped them establish measurable goals to achieve this. The participants ultimately decided to grow small crops in their backyard to sell at the village market days. Once the seeds were planted and sprouted, the team went to visit her garden as the participant took pride in her work and counselors wanted to support her. It was an amazing experience to be able to see the trajectory of this woman’s life change so drastically in such a short amount of time. She went from being depressed, not being able to perform daily activities, and financial stagnation, to a woman with a thriving small crop sales operation.
By the fourth week, attendance at counseling sessions shifted dramatically; previously low turnout was replaced by participants eagerly awaiting our arrival. Word of mouth played a crucial role in this. As participants experienced improvements, their friends began to inquire about the program. Many showed up with friends and family, hoping to enroll. I can recall a day when a mother at Zimmi clinic wished to join the program despite previously not qualifying, as she hadn’t presented with symptoms of depression during her initial screening. This participant explained how since then, she had gotten into an altercation with her partner who eventually abandoned her with their newborn child. She communicated feelings of despair and not knowing who to rely on for guidance. She was screened again and enrolled in the program due to these changes.
To maintain effective communication, weekly recap sessions were held among the counseling team to discuss progress, ways to support one another, tools for sessions, successes, and areas needing improvement.
The MHP included seven one-on-one sessions, with the fifth being a group therapy session. A structured itinerary for the session was used, assigning specific roles to each counselor. For example, a designated counselor led a group prayer, introduced the MHP team, and facilitated an icebreaker through singing. Before discussions, a different counselor explained the session’s flow and encouraged participants to set their own rules, promoting respect and patience. After the group discussion, a counselor closed with prayer and singing to ground everyone before dismissal. New participants who arrived late were welcomed and informed of the rules before joining quietly.
Overall, group therapy sessions were a resounding success, with 24 out of 35 participants (69%) attending. Positive outcomes included participants feeling comfortable sharing their experiences, expressing gratitude for the program, and collaboratively establishing rules that aligned with the session’s purpose.”

The integration phase of the program aimed to provide new or young mothers a path to deal with inciting circumstances associated with PND to prevent relapse and support engagement with the community. Self-help and support groups were initiated by the program for participants who had recovered from their challenges to provide a safe environment to relate to others and maintain daily functioning. Participants took it upon themselves to begin community gardens among other activities that kept them engaged in the community.

#### Initial Training of CBT for Counseling Staff

Counseling staff were trained in CBT curriculum in the field by mental health professionals from MHC-SL. Experts introduced key principles of CBT, emphasizing the creation of a safe space for participants and the importance of self-care skills with a livelihood component. Counselors were taught that their role was not to offer solutions or solve problems, but to help participants develop the skills necessary to do so independently through a gradual process. Below are examples of probing questions that counselors were trained to ask by MHC-SL workers, to understand the key principles of CBT (Table 2).

**Table 2.** Questions used to train CBT counselors.

| **Probing Questions** | **Purpose** |
| --- | --- |
| Since the last time we spoke – How are you doing? | Ice breaking and laying a foundation for onward conversation |
| You sound okay. Am I right to say so? | Allowing participant to dig deep to confirm from an in-depth reflection the status of their wellbeing |
| What has changed so far for the positive? | Appeal to reasoning; assessing how logical is the participant’s flow of thought towards moving away from the past and stop normalizing the abnormal |
| What are existing struggles? | Inquiry is made whether there are still issues to actualizing full recovery of the participant (emotional, social, physical, |
|  | psychological/mental, etc.); working out secondary trauma |
| Do you think that the struggles can be managed? If yes, how? | In depth inquiry is made around reasonable accommodation of cause and effect, while seeking a means to an end |
| What are your next steps? | Digging for alternative reflective thinking to find out whether incidents still cloud thoughts, feelings, and behavior of the participant |

Counseling training began through modules and mock sessions conducted between MHC-SL staff and PPB counselors. Following this, MHC-SL staff observed the initial PPB counseling sessions in person before shifting to a more supportive role.

### Creation of a Depression Screening Tool - The Patient Health Questionnaire (PHQ)

The PHQ is a self-administered diagnostic instrument for common mental disorders. The PHQ-9 specifically assesses depression by scoring each of the nine DSM-IV criteria for depression on a scale from “0” (not at all) to “3” (almost every day). The PHQ-9 has been tested as a reliable and valid measure to both diagnose depressive disorders and determine depression severity (Kroenke et al., 2001). This measure was chosen over a locally developed tool to enhance generalizability by allowing comparison with similar work conducted in different contexts.

### Adapted PHQ-9 (aPHQ-9) for Sierra Leone context

As most measures meant to study psychological distress have been developed for use in other contexts, this study implemented an adapted version of the PHQ-9 and validated it for the use of Sierra Leone. The aim of the adapted tool was to be accepted from a social perspective rather than academic. Modifications were made to the PHQ-9 to exclude culturally insensitive or irrelevant items. The following table describes items that were modified for this study:

All questions also included Mende translations to be carried out in the participants’ native language. The questionnaire put into use was designated as aPHQ-9. aPHQ-9s administration was done by mental health counselors, rather than self-administered. Along with language modifications, the PHQ-9’s used in this study incorporated illustrations of jerry cans to allow participants to visualize the different response options (Figure 2). These illustrations have been tested as a locally validated and culturally appropriate measure of psychological distress in Sierra Leone with good internal consistency (Horn et al., 2021).

**Figure 2.**
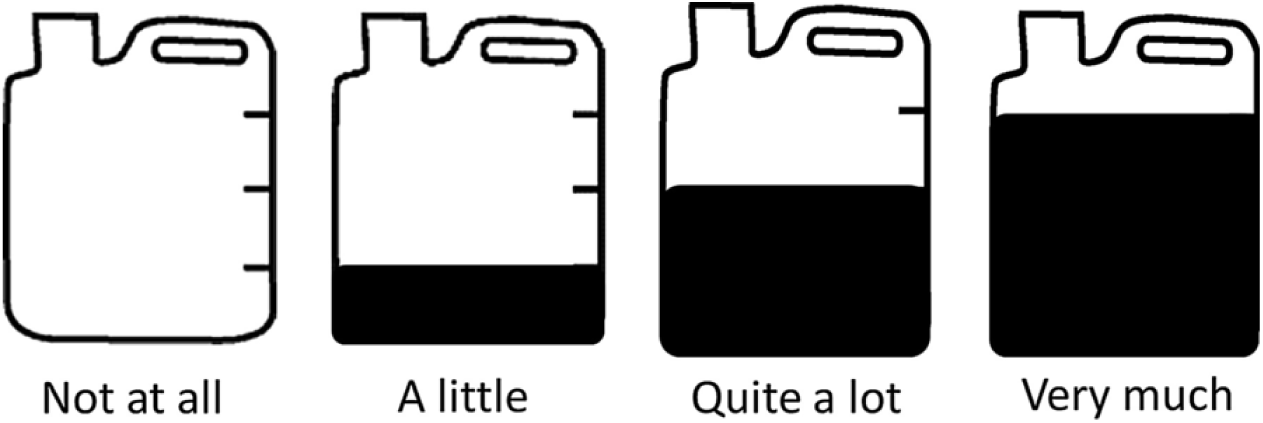
Illustrations of emotion scale response incorporated in PHQ-9 screenings.

This modified version of the PHQ-9 (Figure 3) was successfully field tested in Pujehun District June 2022- October 2022 with >500 women and found to be acceptable and feasible.

**Figure 3.**
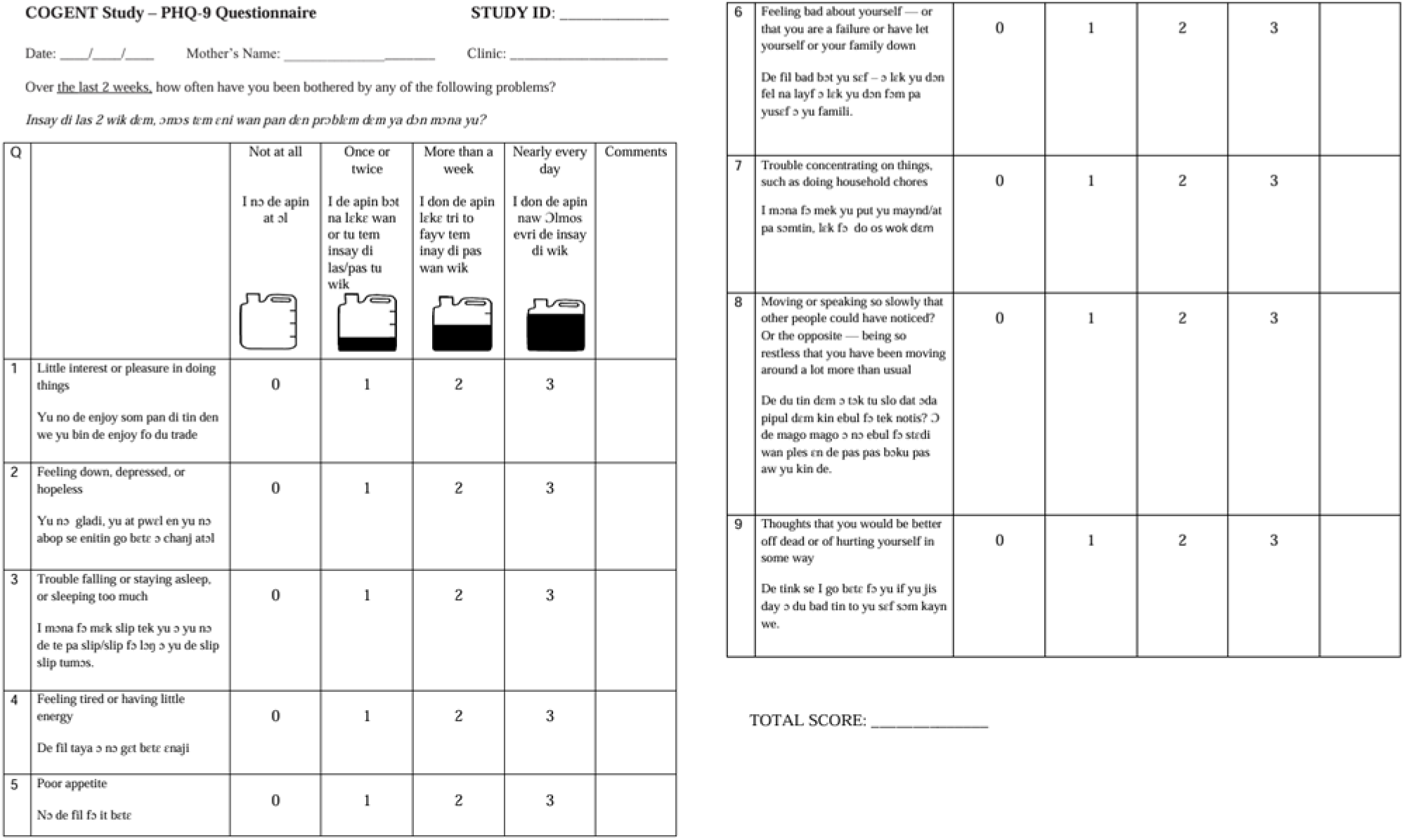
Final aPHQ-9.

### Creation of CBT Curriculum

#### CBT Curriculum

The cut-off score of 9 on the aPHQ-9 was chosen to enroll participants into the CBT program, which has been found to have sensitivity of 90.1% and specificity of 62.0% when diagnosing women with depressive disorder (Gelaye et al., 2013). Once randomized, participants of the CBT study receive an appointment card. The front of the card indicates the participant’s name, study ID number, the antenatal clinic she attends, the date of the initial aPHQ-9 assessment, and the name of the outcome assessor. The inside of the appointment card indicates the date of diagnosis along with the dates of the aPHQ-9 screenings.

The novel CBT program consists of two screening sessions and six 45-60-minute-long sessions conducted once a week. The counselling includes setting goals and/or identifying problems the participant is facing on a weekly basis through a day-to-day plan. Sessions are carried out by the counseling team. While treatment plans are tailored to the participant through collaboration of the counselor and participant, Table 3 is an example of the general structure of CBT sessions.

**Table 3.** PHQ-9 modifications.

| Original Item | Modification |
| --- | --- |
| Item 5: Poor appetite or overeating | Exclude “or overeating” |
| Item 7: Trouble concentrating on things, such as reading the newspaper or watching television | Replace “reading the newspaper or watching television” with “doing household chores” |

While many participants are willing to speak to their counselor after enrollment, two sessions are sometimes necessary to establish a trusting relationship that encourages speaking on personal matters. Intervention techniques include identifying maladaptive thoughts and beliefs, challenging these thoughts and beliefs, behavioral activation, and relaxation. Counseling sessions also involved assigning homework to patients as an effective tool to “facilitate patient skill acquisition, treatment compliance, and symptom reduction by integrating the concepts learned in sessions into daily life” (MHC-SL). Examples of homework assignments included listing obstacles the participant may experience when trying to implement CBT techniques, keeping track of questions regarding CBT that may arise throughout the week, implementing the cognitive-behavioral model for situations that arise throughout the week, and other tailored assignments that counselors deem suitable to re-structure participants’ maladaptive thoughts. For instance, a participant expressing lack of interest in attending school may be assigned to prepare a presentation on a topic of their choice for their next CBT session in hopes of rekindling their interest in learning.

Counselors were aware of the anxiety and allay fears that may come with ending a therapeutic relationship by following an end-of-treatment plan to aid the participant through this process. Ending treatment involves counselors reviewing different cognitive and behavioral skills that the patient has learned throughout their CBT treatment. Measures to prevent relapses are also put in place by anticipating potential stressors the participant may face once treatment has ended and preparing for these inevitable difficulties. Booster sessions are offered to be scheduled approximately 1 month following graduation to check in with the participant and refresh their skills learned in therapy if needed.

The structure of CBT counseling sessions was flexible, and counsellors often worked through multiple problems identified by the participant after successfully covering one. While CBT sessions would ideally be held at the site of enrollment, counselors conduct home visits for participants who are unable to commute to their clinic. All sessions are held in semi-private or private environments. These include a separate room at the clinic, outside of the clinic when a room was unavailable, the participant’s home, or even the car transporting mental health staff when necessary. Sessions also include other parties upon the participants’ request, including relevant family members or spouses.

Eight weeks after enrollment, all participants enrolled in the CBT vs. no CBT study undergo a final aPHQ-9 by their outcome assessor. These scores will then be compared to the baseline scores at enrollment to assess the effectiveness of the novel CBT program.

## Preliminary Results

### Numbers Served

A total of 609 women were counselled through the provisionary mental health service across 5 clinics in Pujehun District, as shown in Table 3. Preliminary field testing of the aPHQ-9 was also conducted to evaluate the feasibility of the tool before launching the formal study. There was a total of 426 participants across 10 clinics in Pujehun District in the aPHQ-9 field testing, as shown in Table 4.

**Table 4.** Structure of CBT curriculum.

|  | **Session Content** |
| --- | --- |
| Session 1 | • Orient participant to CBT<br>• Assess participant's concerns<br>• Set initial treatment plan and goals |
| Session 2 | • Continue assessing participant's concerns as needed<br>• Continue setting goals as needed<br>• Begin intervention techniques |
| Sessions 3-4 | • Begin or continue intervention techniques<br>• Re-assess participant's concerns as needed |
| Session 5 | • Continue intervention techniques<br>• Discuss ending treatment |
| Session 6 | • Continue intervention techniques<br>• Prepare participant to maintain changes post-treatment |

**Table 5.**
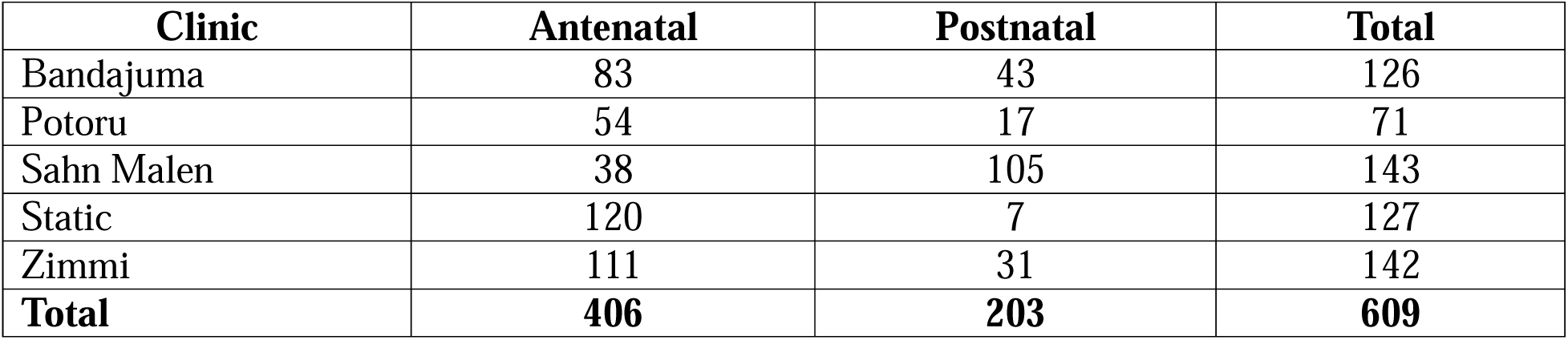
Numbers served by mental health program.

**Table 6.** Numbers served during aPHQ-9 preliminary field testing.

| Clinic | Participants |
| --- | --- |
| Bandajuma | 114 |
| Bendu | 21 |
| Blama Massaquoi | 9 |
| Futa | 8 |
| Gbonapi | 5 |
| Nyandehun | 6 |
| Potoru | 64 |
| Sahn Malen | 34 |
| Static | 66 |
| Zimmi | 99 |
| Total | 426 |

**Table 7.** Overall study design.

| Depression status | CBT vs. none | M-RUSF+ | M-RUSF |
| --- | --- | --- | --- |
| PHQ-9 ≥ 9 | CBT | 1 | 1 |
|  | No CBT | 1 | 1 |
| PHQ-9 < 9 | No CBT |  |  |

### Lessons Learned During Development

During the first two weeks of field testing the aPHQ-9 in Pujehun District, scores were markedly different from previously published PHQ-9 score distributions in previous reports. Based on an epidemiological study conducted in sub-Saharan Africa and studies testing the validity of mental health screening tools in East Africa, up to 34% of participants were expected to be diagnosed with major depressive disorder (Tomlinson et al., 2009; Yotebieng et al., 2024). However, early field testing resulted in a double peaked distribution, with slightly over 50% of participants classified as severely depressed (Figure 3).

Through collaboration with the experts from the MHC-SL, several key observations were made regarding the initial field testing:

1. Several translation modifications were required, mostly for clarification and one significant error

a. The original aPHQ-9 translated “several days,” which corresponds to a score of 1, as “B□ku de d□m,” meaning “very much”

i. This was corrected to the proper Mende translation for “once or twice”
2. Importance of emphasizing symptoms only occurring in the “last 2 weeks” of when screening occurred

a. Participants originally seemed to be recounting any point of their lives at which they may have experienced a particular symptom
3. Need for a comment box to record observations during screening
4. Need for counselors to incorporate many examples when explaining questions to participants

Following these observations and making the appropriate adjustments, score distributions from the aPHQ-9 were much more feasible and in compliance with other PHQ-9 score distributions previously found in Sub-Saharan Africa, with about 5% of participants being diagnosed with moderately severe to severe depression (Figure 4).

**Figure 4.**
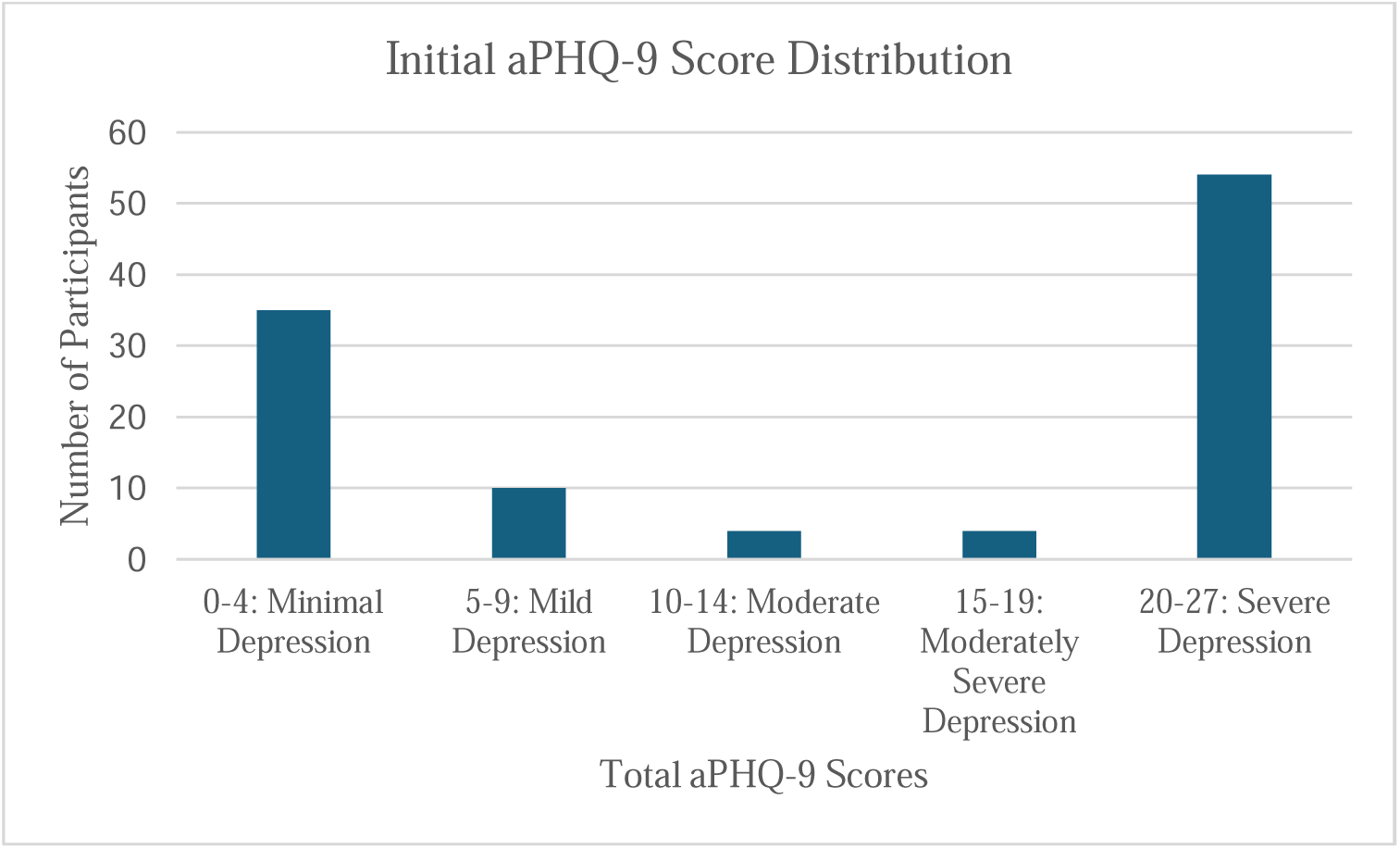
Initial aPHQ-9 score distribution.

**Figure 5.**
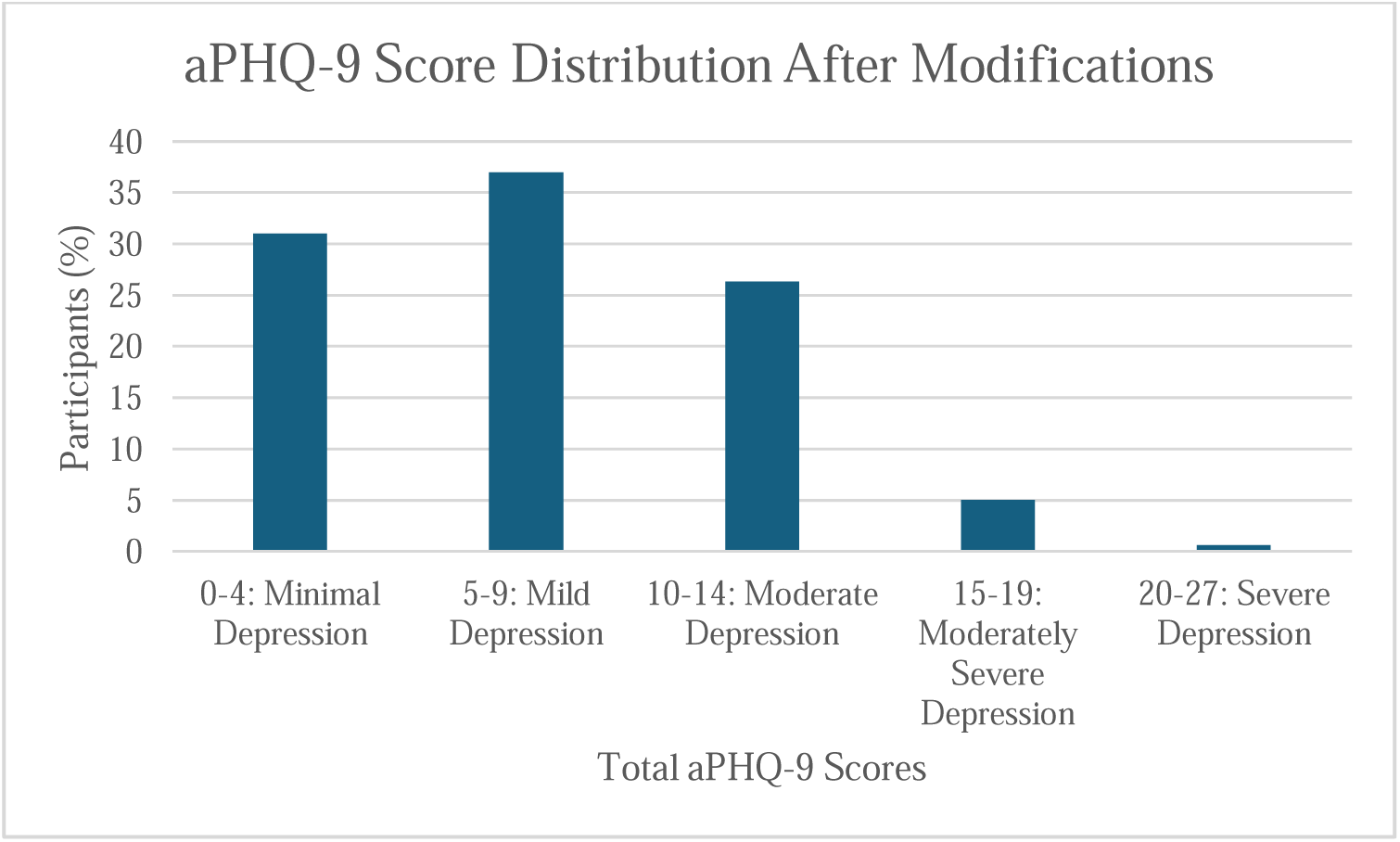
aPHQ-9 score distribution after modifications.

### Overall Response

#### Perspective of Counselors

According to the counselors in Pujehun District, the mental health program has proven to be highly beneficial for many members of the community, and they are proud to be part of a program that has impacted so many lives. They note that some students who have faced abandonment in their homes have become more motivated to continue their education through the support of counseling. Participants whose parents have abandoned them or “left them on the streets” have found a way back to their homes through the program. Many participants also deal with marital and financial problems, which can lead to separation from their spouses. Despite these challenges, there is a strong desire among participants to continue this program, which gives them something they can look forward to, as it has restored hope and transformed their lives.

The following is an excerpt from one of the counselors regarding the impact of PPB’s services, including CBT counselling:

” [Participants’] journey from despair to empowerment illustrates the profound impact of community programs like PPB malnourished pregnant women feeding and CBT. [Their stories serve] as [an] eye opener to the importance of providing support to vulnerable individuals, particularly pregnant women facing challenging circumstances. [Their] resilience and determination after receiving counseling to reclaim […] life are commendable[.]
This report can serve as a foundation for discussions on enhancing more support systems for vulnerable populations in Pujehun District community.”

#### Perspective of Mental Health Coalition

The staff from the MHC-SL believe that this novel mental health counselling program is a necessary service that should continue in the community of Pujehun District and expand to other areas of Sierra Leone. They note an improved quality of life from participants on the program and report positive feedback from participants that see the program and service as a necessity in their lives.

The following case report illustrates the activities of the counseling team.

### Case Report

#### General Information

The participant is a 18 to 23-year-old female who had completed her senior secondary level of education but withdrew from school due to her pregnancy at the time of her initial screening. Her subsequent screening resulted in a score increase from 3 to 10, qualifying her to enroll in the CBT service. Of note, the participant is one of multiple children and was the only child in her family to have pursued higher education.

#### Participant’s CBT

The counselor began the participant’s CBT by asking how her mood and behavior had changed since initial screening. The participant explained that since becoming pregnant, she had moved in with her mother and withdrawn from school. She noted feelings of shame instigated by her mother, who expressed disappointment in her decision to leave school. The participant felt that her family viewed her as a potential source of financial support had she continued her education. During her first session, the participant also conveyed a sense of hopelessness stemming from her mother’s lack of emotional support regarding her pregnancy. The participant mentioned that she had fallen into a routine of arguing with her mother, which would lead her to retreat to her room and cry. After absorbing all this information, the counselor instructed the participant to visualize her ideal scenario. The participant was able to recognize that she would like to re-enroll in school after she gave birth after visualizing her ideal scenario.

The participant acknowledged that much of her distress stemmed from verbal abuse in her home. At the end of her first session, the counselor encouraged her to consider strategies for discussing her plans to return to school with her family.

During the participant’s second CBT session, she shared that her mother refused to listen to her, making it difficult to express her desire to return to school. Nevertheless, the participant reflected more on her plans and recognized that she would need support from her family while raising a baby and pursuing her education. The counselor observed that this marked an improvement from her previous tendency to isolate and cry in her room. When the counselor inquired about other strategies for discussing these issues with her mother, the participant requested that the counselor facilitate the conversation between them.

During the participant’s third session, the counselor facilitated a conversation between her and her mother. The counselor ensured that the participant was able to express herself while stepping in only when necessary. By the end of the discussion, the mother recognized that her actions had caused the participant distress during her pregnancy and promised to help care for the baby, provided the participant returned to school.

#### Techniques employed

- Mood monitoring: counselor asked the participant to reflect on changes in mood since initial enrollment, encouraging self-awareness and self-assessment
- Thought processing: counselor asked the participant about her current feelings regarding her situation, allowing her to recognize and validate these emotions
- Visualization: counselor guided the participant to identify potential goals and reduce feelings of helplessness

`o` Goal setting: through visualization, the participant was able to identify a concrete goal that can help guide her actions and decisions
- Cognitive restructuring: counselor encouraged participant to contrast her current challenges with a hopeful future, resulting in a more positive outlook
- Problem-solving: counselor encouraged participants to consider strategies to promote active problem-solving skills
- Advocacy and communication: counselor facilitated communication between the participant and her family with the participant’s consent, demonstrating participant’s awareness of need for support

## Discussion

### Cultural and Practical Considerations

In partnership with MHC-SL, PPB aimed to develop a counseling program that thrived within the given context. The primary objective was to establish a sustainable process in which women whose lives had diverged with traditional learning due to pregnancy could return to daily activities with tools that helped maintain a productive mindset on their own. This involved considering livelihood needs with an aim to facilitate participants’ reintegration into society. A crucial aspect of developing such a program requires an understanding of depression in the context of rural sub-Saharan Africa and factors that influence the implementation of mental health interventions in this setting.

The MHC-SL advised the use of a community entrance strategy in which PPB addressed key community leaders to introduce the intervention. These community actors included village chiefs, mammy queens, and religious leaders. Gaining support from respected individuals within the community allowed word to spread quickly while also addressing the high level of stigma surrounding mental illness. MHC-SL workers also emphasized the importance of acknowledging the supportive roles of community elders, mostly women who helped care for girls/women affected by pregnancy related problems.

Practical aspects were crucial for implementing this service in Pujehun District. The provisional mental health program acknowledged that participant attendance could be a challenge, particularly during the monsoon season from May to November. To address this, drivers designated junctions that participants commuting from further distances could meet at for pick-up for transportation. Furthermore, counselors made home visits for participants who missed more than one session.

Clinics were held at either Peripheral Health Units (PHU) or Hospitals, with a focus on ensuring privacy. Before starting services, locations were assessed to identify private rooms for counseling. This was done in cooperation with the PHU or hospital staff, to ensure consistency and organization during clinic days. If a PHU lacked adequate privacy, counselors sought alternative solutions such as creating a private space in the PPB vehicle or finding secluded areas nearby. This approach was especially important during house visits, when participants might feel more comfortable discussing sensitive issues away from other members of the household.

PHUs typically designate specific days for perinatal care. CBT clinics were typically scheduled the same day as antenatal care days to maximize attendance. This coordination helped reduce travel time and the burden on women traveling from distant areas by minimizing the number of trips required. Additionally, clinics were aligned with the village’s market days to increase attendance, allowing women to combine their visits to clinic with their trips to market.

Counselors were responsible for record keeping, maintaining detailed notes on attendance and session content after each meeting. This method helped create a supportive environment where participants felt more at ease than if the counselor made observations during the session.

The development and implementation of the counseling service was achieved with $30,000 and served a total of 2114 individuals. The estimated cost of the program development was $17,000, and maintenance of counseling staff was $13,000.

### Future work, a randomized controlled clinical CBT Trial

To determine whether this CBT intervention is truly effective in ameliorating depression, we will evaluate it in a double-blind randomized controlled clinical trial. This will be conducted in the context of the Improving COgnition and GEstational duration with targeted NuTrition (COGENT) study, which studies the gestational duration of malnourished pregnant women receiving two formulations of RUSF, as well as the neurocognitive development of resulting infants (NIH, 2024). This is a 2×2 factorial, randomized, controlled, blinded superiority trial. Women enrolled in the COGENT study will receive monthly depression screenings using the aPHQ-9. Participants of the study will be eligible to enroll in the CBT sub-study at any point of the study if they score a 9 or above on the aPHQ-9.

As depression is a complex condition that is influenced by a variety of factors, including genetic and environmental aspects, diet is thought to play an important role in its etiology and treatment. Diets lacking in essential nutrients have adverse effects on overall brain function and mental health. Dietary fats are a specific class of nutrients associated with mental health, as saturated fats have been correlated with depression while dietary *n*-3 fatty acids are often associated with improvements in depression. One of the essential polyunsaturated fatty acids, α-linoleic acid, is converted to eicosapentaenoic acid (EPA) and docosahexaenoic acid (DHA) which are then incorporated in neurons as functional molecules. It is essential that polyunsaturated fatty acids are consumed through diet, as they are not synthesized endogenously (Fernandes et al., 2017).

A previous randomized controlled trial that evaluated the effect of improved diet quality on symptoms of depression in individuals who met criteria for a major depressive episode and low diet quality found that increased EPA was significantly associated with reduced anxiety and stress at 3 months and 6 months, while increased DHA was associated with reduced stress and negative affect at 6 months (Parletta et al., 2017).

As the intervention RUSF of COGENT was formulated with a fish oil additive containing 500mg of each of the n-3 fatty acids DHA and EPA, in addition to choline, studying this population allows examination of any interactions between n-3 fatty acid deficiency and PND. Table summarizes the overall structure of this study.

The collaboration between PPB and the MHC-SL has successfully established a sustainable counseling program aimed at addressing PND, a critical concern for maternal and infant health that has gone unaddressed in Pujehun District. By engaging community leaders and implementing practical strategies, this initiative empowers women to reintegrate into society, enhancing their well-being and the health outcomes of their children. As we move forward, the upcoming randomized controlled trial within the COGENT study will be vital in assessing the effectiveness of this CBT intervention and its implications for improving both maternal mental health and infant development.

## Data Availability

All data produced in the present study are available upon reasonable request to the authors

## Abbreviations

aPHQ-9: alternative Patient Health Questionnaire
CBT: cognitive behavioral therapy
COGENT: Improving COgnition and GEstational duration with targeted NuTrition
DHA: Docosahexaenoic acid
EPA: Eicosapentaenoic acid
MHC-SL: Mental Health Coalition
MoHS: Ministry of Health and Sanitation
PHU: Peripheral Health Unit
PND: perinatal depression
PPB: Project Peanut Butter
RUSF: Ready to Use Supplemental Food
RUTF: Ready to Use Therapeutic Food
SMART: Specific, Measurable, Attainable, Realistic, Timely
SSA: sub-Saharan Africa

## References

Dagher, R. K., Bruckheim, H. E., Colpe, L. J., Edwards, E., & White, D. B. (2021). Perinatal Depression: Challenges and Opportunities. Journal of Women’s Health, 30(2). 10.1089/jwh.2020.8862

Endomba, F. T., Ndoadoumgue, A. L., Mbanga, C. M., Nkeck, J. R., Ayissi, G., Danwang, C., & Bigna, J. J. (2021). Perinatal depressive disorder prevalence in Africa: A systematic review and Bayesian analysis. General Hospital Psychiatry, 69, 55–60. 10.1016/j.genhosppsych.2021.01.006

Fernandes, M., Mutch, D., & Leri, F. (2017). The Relationship between Fatty Acids and Different Depression-Related Brain Regions, and Their Potential Role as Biomarkers of Response to Antidepressants. Nutrients, 9(3), 298. 10.3390/nu9030298

Gelaye, B., Williams, M. A., Lemma, S., Deyessa, N., Bahretibeb, Y., Shibre, T., Wondimagegn, D., Lemenhe, A., Fann, J. R., Vander Stoep, A., & Andrew Zhou, X.-H. (2013). Validity of the patient health questionnaire-9 for depression screening and diagnosis in East Africa. Psychiatry Research, 210(2), 653–661. 10.1016/j.psychres.2013.07.015

Hofmann, S. G., Asnaani, A., Vonk, I. J. J., Sawyer, A. T., & Fang, A. (2012). The Efficacy of Cognitive Behavioral Therapy: a Review of Meta-Analyses. Cognitive Therapy and Research, 36(5), 427–440. 10.1007/s10608-012-9476-1

Horn, R., Jailobaeva, K., Arakelyan, S., & Ager, A. (2021). The development of a contextually appropriate measure of psychological distress in Sierra Leone. BMC Psychology, 9(1). 10.1186/s40359-021-00610-w

Hunn, L., Teague, B., & Fisher, P. (2023). Literacy and Mental Health Across the Globe: A Systematic Review. Mental Health and Social Inclusion, 27(4). 10.1108/mhsi-09-2022-0064

Kroenke, K., Spitzer, R. L., & Williams, J. B. W. (2001). The PHQ-9: Validity of a brief depression severity measure. Journal of General Internal Medicine, 16(9), 606–613. 10.1046/j.1525-1497.2001.016009606.x

Lane, R. I. (2021). “We Are Here,” Enduring Survivorship In The Aftermath: Community Trauma, Survivor Discourse and Advocacy Post the 2014-16 West Africa Ebola Outbreak. Escholarship.org. https://escholarship.org/uc/item/1pw1c1vb

M’Cormack-Hale, F., & Akua Amoah Twum, M. (2022). Gender-based violence a high priority in Sierra Leone, but citizens say it is a private matter. In Afrobarometer. https://www.afrobarometer.org/wp-content/uploads/2022/10/AD565-Gender-based-violence-a-top-priority-in-Sierra-Leone-Afrobarometer-25oct22-1.pdf

Mayo Clinic. (2019, March 16). Cognitive Behavioral Therapy. Mayoclinic.org; Mayo Clinic. https://www.mayoclinic.org/tests-procedures/cognitive-behavioral-therapy/about/pac-20384610

Mental Health Innovation Network. (2014). Mental Health Coalition – Sierra Leone | MHIN. MHIN. https://www.mhinnovation.net/innovations/mental-health-coalition-sierra-leone

NIH. (2024). Improving Cognition and Gestational Duration With Targeted Nutrition (COGENT). Clinicaltrials.gov. https://clinicaltrials.gov/study/NCT05949190

O’Connor, E., Senger, C. A., Henninger, M., Gaynes, B. N., Coppola, E., & Weyrich, M. S. (2019, February). Introduction. Nih.gov; Agency for Healthcare Research and Quality (US). https://www.ncbi.nlm.nih.gov/books/NBK537822/#ch1.s4

Parletta, N., Zarnowiecki, D., Cho, J., Wilson, A., Bogomolova, S., Villani, A., Itsiopoulos, C., Niyonsenga, T., Blunden, S., Meyer, B., Segal, L., Baune, B. T., & O’Dea, K. (2017). A Mediterranean-style dietary intervention supplemented with fish oil improves diet quality and mental health in people with depression: A randomized controlled trial (HELFIMED). Nutritional Neuroscience, 22(7), 474–487. 10.1080/1028415x.2017.1411320

Psaltopoulou, T., Sergentanis, T. N., Panagiotakos, D. B., Sergentanis, I. N., Kosti, R., & Scarmeas, N. (2013). Mediterranean diet, stroke, cognitive impairment, and depression: A meta-analysis. Annals of Neurology, 74(4), 580–591. 10.1002/ana.23944

Rathod, S., Phiri, P., & Naeem, F. (2019). An evidence-based framework to culturally adapt cognitive behaviour therapy. The Cognitive Behaviour Therapist, 12(12). 10.1017/s1754470x18000247

ReliefWeb. (2016, January 4). Sierra Leone: Pujehun District Profile (04 December 2015) - Sierra Leone. ReliefWeb. https://reliefweb.int/report/sierra-leone/sierra-leone-pujehun-district-profile-04-december-2015

Tomlinson, M., Grimsrud, A., Stein, D., Williams, D., & Myer, L. (2009). The epidemiology of major depression in South Africa: Results from the South African Stress and Health study. South African Medical Journal, 99(5). https://scholar.harvard.edu/sites/scholar.harvard.edu/files/davidrwilliams/files/2009-the_epidemiology_of-williams.pdf

World Health Organization. (2017). Depression and Other Common Mental Disorders Global Health Estimates. https://iris.who.int/bitstream/handle/10665/254610/W?sequence=1

World Health Organization. (2023). Depressive disorder (depression). World Health Organization; World Health Organization. https://www.who.int/news-room/fact-sheets/detail/depression

World Population Review. (2024, November 4). Sierra Leone Population 2024 (Live). Worldpopulationreview.com. https://worldpopulationreview.com/countries/sierra-leone

Yotebieng, M., Zotova, N., Bernard, C., Goodrich, S., Ajeh Rogers Awoh, Watnick, D., Nsonde, D. M., Flore, E., Laure, J., Calvin, G., Minga, A., Moussa Seydi, Gandou, P., Kwobah, E. K., Lukoye Atwoli, Jaquet, A., Wools-Kaloustian, K., & Anastos, K. (2024). Accuracy of PHQ-9 against psychiatric diagnosis for depression among people living with HIV: A multicounty cross-sectional study. AIDS. 10.1097/qad.0000000000003963

